# Examining Perinatal Regionalization in Practice: A Network Analysis of Maternal Transport in Georgia

**DOI:** 10.1101/2025.02.14.25322280

**Authors:** Jingyu Li, Stephanie M. Radke, Lauren N. Steimle

**Affiliations:** H. Milton Stewart School of Industrial and Systems Engineering, Georgia Institute of Technology, Atlanta, GA, USA; Department of Obstetrics & Gynecology, University of Iowa Hospitals & Clinics, Iowa City, Iowa, USA

## Abstract

**Objective:** The primary objective was to use network analysis to characterize maternal transport patterns in the state of Georgia and compare them with the state’s designated perinatal regions (DPRs).

**Study Design:** Using 2017-2022 birth records in Georgia, we constructed network graphs of maternal transport routes among obstetric facilities. We used multivariate logistic regression to identify factors associated with inter-DPR transports. We applied a community-detection algorithm to cluster facilities and compared the clusters to Georgia’s DPRs.

**Results:** Among 774 639 deliveries, 2 757 (0.36%) involved transports among obstetric facilities. 8 facility clusters were identified and strongly aligned with DPRs (p < 0.001). Inter-DRP transports tended to occur between neighboring DPRs and between facilities belonging to the same healthcare system (p < 0.001).

**Conclusion:** Network analysis reveals patterns of maternal transports among obstetric facilities. States can improve the design of perinatal regionalization systems by formalizing existing partnership among obstetric facilities.

## Introduction

Maternal mortality in the United States (U.S.) is rising and among the highest in developed countries, with significant disparities across racial, ethnic, and socio-economic groups.^1,2^ Rural populations are particularly affected, with noncore areas and micropolitan areas experiencing 37.9 and 31.2 pregnancy-related deaths per 100 000 live births in 2020, respectively.^3^ Geographic barriers and a lack of access to obstetric care in rural regions are linked to higher maternal morbidities and mortality.^4–6^

In response, the American College of Obstetricians and Gynecologists has emphasized the need to improve the delivery of maternal care at a systems-level. Perinatal regionalization, also known as “risk-appropriate care,” is a systems-level strategy for coordinating care to ensure pregnant people receive timely care at facilities with risk-appropriate personnel and services.

The strategy guides states to develop coordinated systems that designate the *levels of care* of each facility to reflect which types of pregnancies the facility is suited to handle.^7^ However, perinatal regionalization has historically focused on neonatal care and formal maternal regionalization systems are only recently emerging.^8–10^

Due to varying capabilities of facilities to provide different levels of maternal care, inter-facility maternal transport is a crucial component of the risk-appropriate care. When a facility cannot provide an appropriate level of care for a complicated pregnancy, prompt transfer to a higher-level facility can decrease maternal, fetal, and neonatal morbidity and mortality.^11,12^ Improvements to perinatal regionalization, especially the coordination of inter-facility maternal transport, present opportunities to prevent pregnancy-related deaths.^11^ Inter-facility transport policies are crucial to enhance care coordination during an obstetric emergency, particularly in rural regions.^13^ Within these regionalized systems, higher level facilities are expected to provide training and education to facilities that refer to them. As of 2019, 39 states had established inter-facility transport policies and 37 states had policies with specific protocols for maternal transport.^14^

Despite recent recognition of maternal regionalization systems, the implementation and adherence to state-specific guidelines for maternal transport can vary significantly.^15^ Inconsistent practices can lead to delays or failures in receiving specialized care, which may lead to life-threatening complications.^11^ A recent study of seven geographically diverse states showed that 43% of high-risk pregnancies occurred in facilities that were not risk-appropriate.^16^ The evaluation of maternal regionalization systems and their alignment with state guidelines is urgently needed to identify barriers to implementation. In this study, we characterized maternal transports in the state of Georgia using network analysis, which has emerged as a powerful data-driven approach in multiple public health contexts to examine perinatal regionalization systems in practice.^17–19^ The purpose of this study was to characterize deliveries involving maternal transports, construct maternal transport networks to represent transport routes among obstetric facilities, characterize maternal transports across the state’s designated perinatal regions (DPRs) and compare observed maternal transport networks with the state’s guidelines.

## Materials and Methods

### Overview

Since 1970s, obstetric facilities in Georgia started adopting perinatal level designations to establish a regionalized system of perinatal care. In 2017, the Georgia Department of Public Health updated its guidelines to align with current contract standards and improve the delivery of risk-appropriate maternal care through coordination among facilities with different levels of care.^20^ Obstetric facilities in Georgia were categorized into three levels of care: Basic (Level I), Specialty (Level II), and Subspecialty (Level III). While some facilities were practicing according to Level IV standards, none were officially designated as such in Georgia as of 2017. These guidelines strategically partitioned the state’s counties into six DPRs: Albany, Atlanta, Augusta, Columbus, Macon, and Savannah. Each DPR has at least one designated Regional Perinatal Center (RPC), which offers the highest level of care to serve a defined geographic region and coordinate care within their region.

In this analysis, we first compared profiles of pregnant people in the transport and non-transport groups. We also compared the rates of transports by rurality of the county of residence. To examine regionalization of maternal transport in Georgia, we constructed *maternal transport networks* which capture the flow and distribution of patient transport across different facilities.

### Study sample

We used birth records from the state of Georgia between January 1^st^, 2017 and December 31^st^, 2022. To construct maternal transport networks, we excluded deliveries in which the pregnant people was not transported and those with missing origin or destination facility records. We also excluded transports from or to non-obstetric facilities. Obstetric facilities were a list of facilities with obstetric units which were included in the 2017 Georgia guidelines for perinatal regionalization.^20^ We focused on transports among those facilities since care coordination was only defined for obstetric facilities under previous state guidelines. **Figure 1** provides more details about the data inclusion criteria for included samples. The [Blinded for Peer Review] Institutional Review Board approved this study (Protocol H23091). Description of variables analyzed in this study is detailed in **Appendix Table A1**.

**Figure 1.**
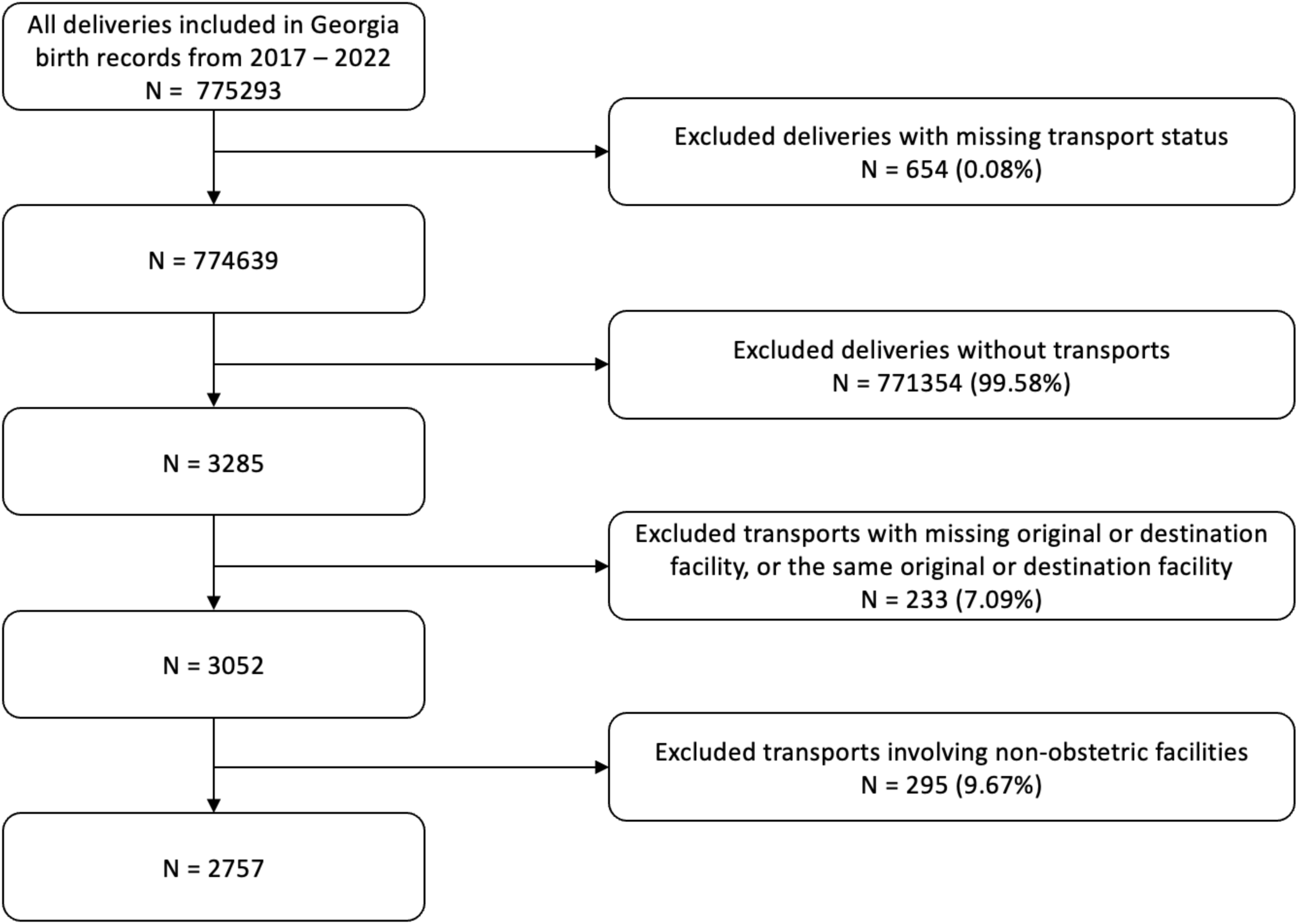
Data inclusion criteria for the study sample. To construct maternal transport networks among obstetric facilities, we excluded deliveries that had missing transport status, did not involve a transport, had missing records of original or destination facility, had the same original or destination facility, or involved non-obstetric facilities.

### Objective 1: Characterization of transported deliveries

We partitioned our study sample into deliveries with and without maternal transport and compared demographic information, medical risk factors, and other pregnancy-related conditions between these groups. We used unadjusted odds ratios to assess statistical significance. We reported maternal transport rates per 1 000 resident births by county of residence, defined as the total number of transports among people who live in that county per 1 000 births to people who live in that county.

### Objective 2: Representation of maternal transport networks

We constructed maternal transport networks using obstetric facilities included in the Georgia’s 2017 guidelines. For each transport, we identified the *origin* (the facility transferring the patient), and the *destination* (where the birth occurred). Then, we constructed a directional network graph to represent maternal transports from the origin to the destination (see **Figure 2**). In the graph, nodes represent unique obstetric facilities and directed edges represent at least one transport from the origin to the destination. We assigned edge weights to represent the number of transports between facilities.

**Figure 2.**
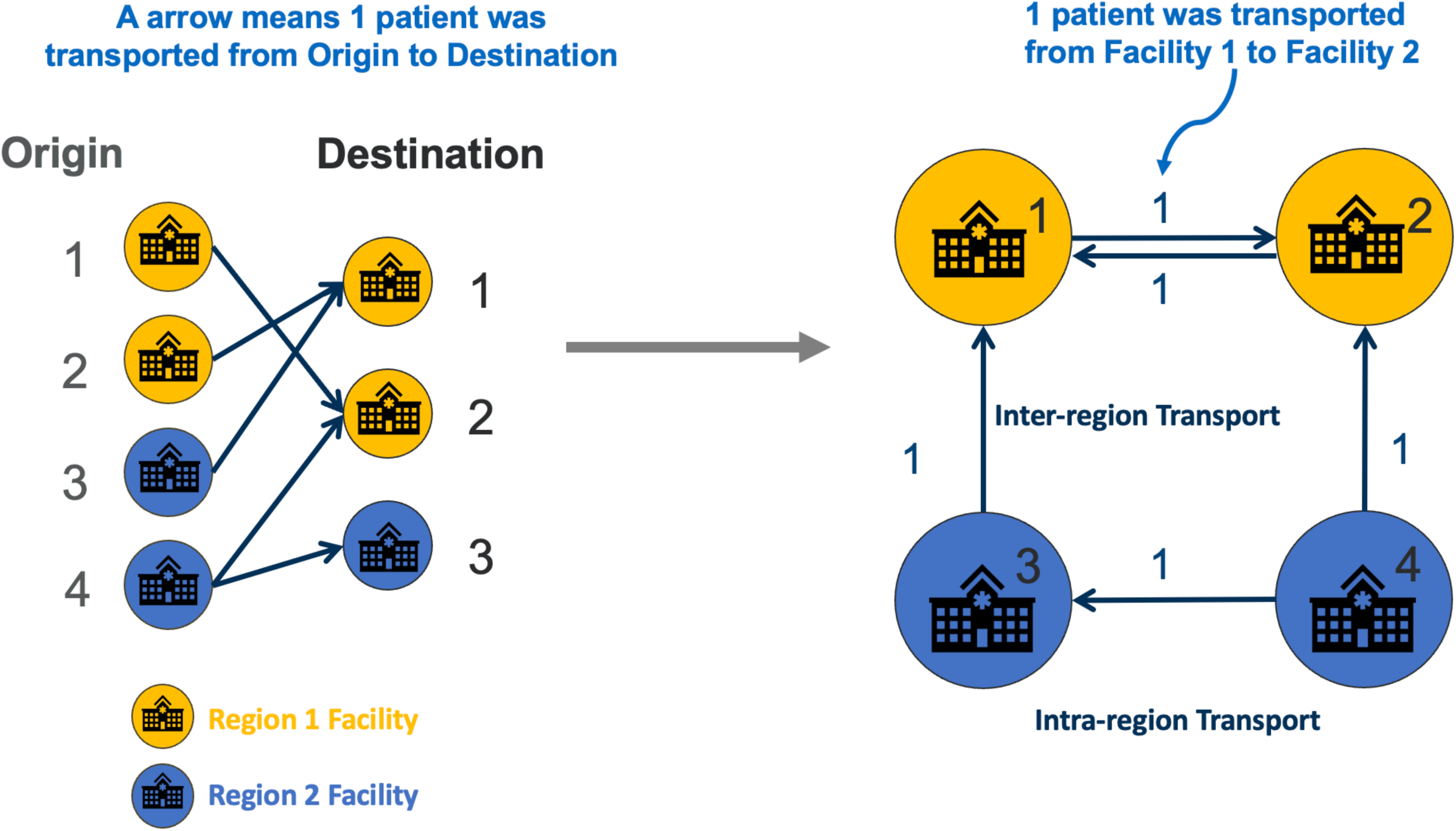
Illustration of the network representation of maternal transport routes. Each node (circle) indicates a facility with its color representing its designated perinatal region. Arrows represent that a transport occurred between the indicated origin facility and the destination facility.

### Objective 3: Characterization of the inter-region maternal transport

We characterized the extent to which facilities transport people across DPRs. We labeled a transport as an “inter-region transport” if the origin and the destination facilities were in different DPRs and as an “intra-region transport” otherwise. We derived the percentage of nodes that transported patients from or to nodes from another DPR, the percentage of edges that connected nodes from another DPR, and the percentage of transport volumes that flow between DPRs. To examine factors associated with inter-region transport, we used a multivariate logistic regression model. We included both patient-level factors (demographics, location, medical risks) and facility-level factors (level of care, healthcare systems) to predict whether the origin facility and destination facility are across DPRs.

### Objective 4: Comparison of observed networks and state guidelines for regionalization

We characterized “communities” of facilities within the network among which transports occurred the most frequently. “Community detection” is a process of identifying groups of nodes in a network that are more densely connected to each other while minimizing number of connections between groups. In the context of maternal transport, algorithm-detected communities represent groups of facilities that an algorithm determines are most likely to transport amongst themselves. See **Appendix A3** for more technical details.

The multivariate logistic regression model was built using R (version 4.3.3). All other analyses were performed in Python (version 3.11.7). The network analysis was conducted using NetworkX package.^21^ We used a p-value of 0.05 as the significance threshold throughout this analysis.

## Results

### Study sample

The study included 774 639 deliveries, with 3 285 (0.42%) involving a maternal transport (see **Table 1**), among which 2 757 (0.36%) occurred between obstetric facilities. Among all included deliveries (mean [SD] maternal age: 28.6 [5.89] years), 81.5% of the deliveries were to urban residents, 11.8% to suburban residents, and 4.0% to rural residents. 56.7% were to White people, 35.8% to Black or African-American people, 4.6% to Asian people, 2.6% to multiracial people, 0.2% to Native Hawaiian or Pacific Islanders, and 0.1% to American Indian or Alaska Native people. 3.9% of these deliveries had a breech presentation, and 34.4% were Cesarean deliveries.

**Table 1.** Characteristics of the study sample by transport status.

|  | **Non-transport**<br>**(n = 771354)** | **Transport**<br>**(n = 3285)** | **Total**<br>**(n = 774639)** | **Unadjusted OR (95% CI)** | **p value** |
| --- | --- | --- | --- | --- | --- |
| **Age in years, mean (SD)** | 28.6 (5.89) | 27.5 (5.93) | 28.6 (5.89) | 0.968 (0.962-0.974) | <0.001*** |
| **Race, n (%)** |  |  |  |  |  |
| American Indian or Alaska Native | 1042 (0.1%) | 3 (0.1%) | 1045 (0.1%) | 0.823 (0.265-2.558) | 0.736 |
| Asian | 35869 (4.7%) | 40 (1.2%) | 35909 (4.6%) | 0.319 (0.233-0.436) | <0.001*** |
| Black or African-American | 275801 (35.8%) | 1617 (49.2%) | 277418 (35.8%) | 1.675 (1.562-1.797) | <0.001*** |
| Multiracial | 19683 (2.6%) | 89 (2.7%) | 19772 (2.6%) | 1.292 (1.043-1.601) | 0.019* |
| Native Hawaiian or Other Pacific Islander | 1487 (0.2%) | 5 (0.2%) | 1492 (0.2%) | 0.961 (0.399-2.315) | 0.929 |
| White | 437472 (56.7%) | 1531 (46.6%) | 439003 (56.7%) | ref |  |
| **Ethnicity, n (%)** |  |  |  |  |  |
| Hispanic | 114056 (14.8%) | 258 (7.9%) | 114314 (14.8%) | ref |  |
| Non-Hispanic | 652216 (84.6%) | 2988 (91.0%) | 655204 (84.6%) | 2.025 (1.783-2.300) | <0.001*** |
| Unknown | 5082 (0.7%) | 39 (1.2%) | 5121 (0.7%) | 3.393 (2.420-4.756) | <0.001*** |
| **Location of residence<sup>a</sup>, n (%)** |  |  |  |  |  |
| Urban | 629466 (81.6%) | 1573 (47.9%) | 631039 (81.5%) | ref |  |
| Suburban | 90063 (11.7%) | 1158 (35.3%) | 91221 (11.8%) | 5.145 (4.768-5.553) | <0.001*** |
| Rural | 30514 (4.0%) | 431 (13.1%) | 30945 (4.0%) | 5.652 (5.078-6.292) | <0.001*** |
| Unknown | 21311 (2.8%) | 123 (3.7%) | 21434 (2.8%) | 2.310 (1.921-2.776) | <0.001*** |
| **Education Level, n (%)** |  |  |  |  |  |
| Less than 9<sup>th</sup> grade | 24293 (3.1%) | 85 (2.6%) | 24378 (3.1%) | 0.977 (0.785-1.216) | 0.835 |
| 9th through 11<sup>th</sup> grade | 68362 (8.9%) | 424 (12.9%) | 68786 (8.9%) | 1.732 (1.555-1.929) | <0.001*** |
| high school diploma or GED | 244075 (31.6%) | 1216 (37%) | 245291 (31.7%) | 1.391 (1.290-1.500) | <0.001*** |
| Some college or higher | 431393 (55.9%) | 1545 (47.0%) | 432938 (55.9%) | ref | <0.001*** |
| Unknown | 3231 (0.4%) | 15 (0.5%) | 3246 (0.4%) | 1.296 (0.779-2.158) | 0.318 |
| **Payor, n (%)** |  |  |  |  |  |
| Commercial Insurance | 311,055 (40.3%) | 738 (22.5%) | 311,793 (40.3%) | ref |  |
| Medicaid, Medicaid Applicants, or Medicaid Managed Care | 353,994 (45.9%) | 2026 (61.7%) | 356020 (46.0%) | 2.412 (2.217-2.625) | <0.001*** |
| Other Insurance, Government Assistance, or Other Non-specified Managed Care | 25,528 (3.3%) | 122 (3.7%) | 25650 (3.3%) | 2.014 (1.662-2.441) | <0.001*** |
| Self-Pay | 49289 (6.4%) | 123 (3.7%) | 49412 (6.4%) | 1.052 (0.869-1.273) | 0.604 |
| TRICARE | 30385 (3.9%) | 274 (8.3%) | 30659 (4.0%) | 3.801 (3.307-4.368) | <0.001*** |
| Unknown | 1103 (0.1%) | 2 (0.1%) | 1105 (0.1%) | 0.764 (0.191-3.062) | 0.704 |
| **APNCU Index<sup>b</sup>, n (%)** |  |  |  |  |  |
| Inadequate | 126318 (16.4%) | 577 (17.6%) | 126895 (16.4%) | ref |  |
| Intermediate | 51106 (6.6%) | 155 (4.7%) | 51261 (6.6%) | 0.664 (0.556-0.793) | <0.001*** |
| Adequate | 245791 (31.9%) | 529 (16.1%) | 246320 (31.8%) | 0.471 (0.419-0.530) | <0.001*** |
| Adequate Plus | 302521 (39.2%) | 1758 (53.5%) | 304279 (39.3%) | 1.272 (1.158-1.398) | <0.001*** |
| Unknown | 45618 (5.9%) | 266 (8.1%) | 45884 (5.9%) | 1.277 (1.104-1.477) | 0.001** |
| **Tobacco use, n (%)** |  |  |  |  |  |
| Yes | 30104 (3.9%) | 320 (9.7%) | 30424 (3.9%) | 2.663 (2.371-2.990) | <0.001*** |
| No | 737690 (95.6%) | 2945 (89.7%) | 740635 (95.6%) | ref |  |
| Unknown | 3560 (0.5%) | 20 (0.6%) | 3580 (0.5%) | 1.407 (0.905-2.187) | 0.129 |
| **Number of Medical Risk Factors<sup>c</sup>, n (%)** |  |  |  |  |  |
| 0 | 554204 (71.8%) | 1587 (48.3%) | 555791 (71.7%) | ref |  |
| 1 | 175999 (22.8%) | 1118 (34.0%) | 177117 (22.9%) | 2.218 (2.055-2.395) | <0.001*** |
| >=2 | 39268 (5.09%) | 553 (16.83%) | 39821 (5.14%) | 4.918 (4.462-5.421) | <0.001*** |
| Unknown | 1883 (0.2%) | 27 (0.8%) | 1910 (0.2%) | 5.007 (3.414-7.345) | <0.001*** |
| **Gestation Weeks<sup>d</sup>, n (%)** |  |  |  |  |  |
| < 37 weeks | 87860 (11.4%) | 2529 (77.0%) | 90389 (11.7%) | 26.057 (24.016-28.272) | <0.001*** |
| >= 37 weeks | 683461 (88.6%) | 755 (23.0%) | 684216 (88.3%) | ref |  |
| Unknown | 33 (0.0%) | 1 (0.0%) | 34 (0.0%) | 27.432 (3.747-200.82) | 0.001** |
| **Plurality, n (%)** |  |  |  |  |  |
| 1 | 744751 (96.6%) | 2783 (84.7%) | 747534 (96.5%) | ref |  |
| >=2 | 26603 (0.4%) | 502 (15.3%) | 27105 (0.5%) | 5.050 (4.588-5.558) | <0.001*** |
| **Fetal Presentation, n (%)** |  |  |  |  |  |
| Breech | 29400 (3.8%) | 544 (16.6%) | 29944 (3.9%) | 5.074 (4.624-5.568) | <0.001*** |
| Cephalic | 734342 (95.2%) | 2678 (81.5%) | 737020 (95.1%) | ref |  |
| Other | 5981 (0.8%) | 58 (1.8%) | 6039 (0.8%) | 2.659 (2.048-3.453) | <0.001*** |
| Unknown | 1631 (0.2%) | 5 (0.2%) | 1636 (0.2%) | 0.841 (0.349-2.024) | <0.001*** |
| **Final Method of Delivery, n (%)** |  |  |  |  |  |
| Cesarean | 264821 (34.3%) | 1879 (57.2%) | 266700 (34.4%) | 2.557 (2.386-2.740) | <0.001*** |
| Vaginal | 505927 (65.6%) | 1404 (42.7%) | 507331 (65.5%) | ref |  |
| Unknown | 606 (0.1%) | 2 (0.1%) | 608 (2.1%) | 1.189 (0.296-4.771) | 0.807 |
| **Infant Transfer after Birth, n (%)** |  |  |  |  |  |
| Yes | 6267 (0.8%) | 40 (1.2%) | 6307 (0.8%) | 1.505 (1.101-2.057) | 0.011* |
| No | 764074 (99.1%) | 3241 (98.7%) | 767315 (99.1%) | ref |  |
| Unknown | 1013 (0.1%) | 4 (0.1%) | 1017 (0.1%) | 0.931 (0.349-2.487) | 0.886 |
| **Abnormal Condition relating to NICU Admission, n (%)** |  |  |  |  |  |
| Yes | 71909 (9.3%) | 2302 (70.1%) | 6307 (0.8%) | 22.778 (21.131-24.554) | <0.001*** |
| No | 699444 (90.7%) | 983 (29.9%) | 767315 (99.1%) | ref |  |
| Unknown | 1 (0.0%) | 0 (0.0%) | 1017 (0.1%) | 0.018 (0-inf) | 0.973 |

Abbreviations: OR, odds ratio; CI, confidence interval; SD, standard deviation; ref, reference level; GED, general education development.
- Location of residence: we categorized patients' residence by linking 2020 census block group to the Rural-Urban Continuum Codes (RUCCs): urban (RUCCs of 1-3), suburban (RUCCs of 4-6), and rural (RUCCs of 7-9). - APNCU index: the Adequacy of Prenatal Care Utilization Index. This index determines the adequacy of prenatal care utilization based on two parts: the month in which prenatal care is initiated and the number of visits from initiation of care until delivery. It is categorized into four categories: inadequate, intermediate, adequate, and adequate plus. - Medical risk factors: pre-pregnancy diabetes, gestational diabetes, eclampsia, pre-pregnancy hypertension, gestational/pregnancy-associated hypertension, previous preterm birth, and previous Cesarean delivery. - Preterm birth refers to any deliveries with gestation weeks less than 37 weeks.
\*: significant at p=0.05; \*\*: significant at p=0.01; \*\*\*: significant at p=0.001

### Objective 1: Characterization of transported deliveries

Significant differences were observed between the demographics in the non-transport and the transport groups (see **Table 1**). The transport group was more likely to include those who were Black or African-American (unadjusted OR: 1.675, 95% CI: 1.562-1.797, p < 0.001, ref: White), living in an rural (unadjusted OR: 5.145, 95% CI: 4.768-5.553, p < 0.001, ref: urban residence) or suburban area (unadjusted OR: 5.652, 95% CI: 5.078-6.292, p < 0.001, ref: urban residence), and being Medicaid-insured (unadjusted OR: 2.412, 95% CI: 2.217-2.625, p < 0.001, ref: commercially insured) or TRICARE-insured (unadjusted OR: 3.801, 95% CI: 3.307-4.368, P < 0.001, ref: commercially insured).

Several behavioral and medical risk factors were also significantly associated with maternal transports (**Table 1**), such as tobacco use (unadjusted OR: 2.663, 95% CI: 2.371-2.990, p < 0.001, ref: no tobacco use) and having multiple medical conditions (unadjusted OR: 4.918, 95% CI: 4.462-5.421, p < 0.001, ref: no reported medical conditions). Characteristics related to complicated deliveries, including preterm delivery (unadjusted OR: 26.057, 95% CI: 24.016-28.272, p < 0.001), multiple births (unadjusted OR: 5.050, 95% CI: 4.588-5.558, p < 0.001), subsequent infant transfer (unadjusted OR: 1.505, 95% CI: 1.101-2.057, p = 0.011), and admission to NICU due to abnormal conditions (unadjusted OR: 22.778, 95% CI: 21.131-24.554, p < 0.001) were also associated with maternal transport.

**Figure 3** illustrates the maternal transport rate by county (mean [SD]: 9.35 [8.62] transports per 1 000 resident births per county). We observed that counties in the southern Macon region and the southwestern part of the Albany region exhibited the highest rates of maternal transport.

**Figure 3.**
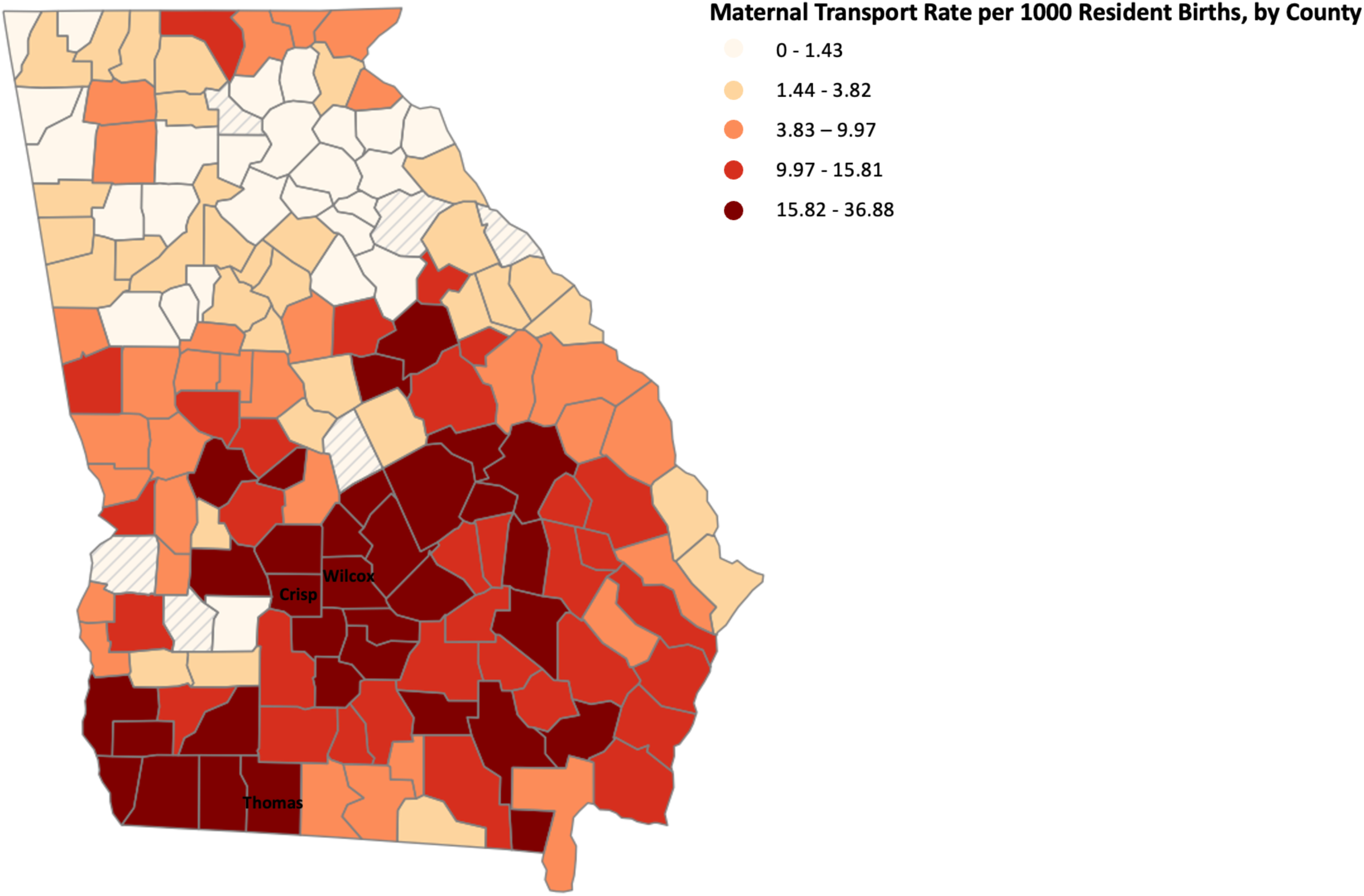
Maternal transport rates by county of residence for pregnant people (mean [SD]: 9.35 [8.62] transports per 1 000 resident births). Wilcox County has the highest maternal transport rate of 36.88 transports per 1 000 resident births, followed by Thomas County of 31.62 transports per 1 000 resident births and Crisp County of 31.11 transports per 1 000 resident births. Maternal transport rates were defined as the total number of transports among people who lived in that county per 1 000 births to people who lived in that county. Hashed lines represent counties with zero transport. Rates were divided into 5 quantiles, indicated by the shade of colorings. The mean maternal transport rate was calculated by taking the transport rate of each county divided by the total number of counties in Georgia.

Out of 159 counties in Georgia, Wilcox County has the highest maternal transport rate of 36.88 transports per 1 000 resident births, followed by Thomas County of 31.62 transports per 1 000 resident births and Crisp County of 31.11 transports per 1000 resident births. We also observed that the average maternal transport rates among urban counties (mean [SD]: 4.75 [4.99] per 1 000 resident births) were much lower than the average maternal transport rates among suburban (mean [SD]: 13.34 [9.41] per 1 000 resident births) and rural counties (mean [SD]: 13.37 [8.87] per 1 000 resident births).

### Objective 2: Representation of maternal transport networks

After removing deliveries with incomplete origin or destination facilities and excluding non-obstetric facilities, 2 757 transports remained across 199 unique transport routes (i.e., unique origin-destination pairs), which were used in subsequent analyses. **Table 2** shows the number of transports by levels of care of the origin and destination facilities. 2 629 (95%) of transports were from lower-level facilities to higher-level facilities. Among those, 2 289 (87%) were to an RPC.

**Table 2.** Maternal transport by levels of care.

|  | *Destination* |  |  |  |
| --- | --- | --- | --- | --- |
|  | Level I | Level II | Level III | RPC |
| ***Origin*** |  |  |  |  |
| Birth Center | 0 | 0 | 223 (79.6%) | 57 (20.4%) |
| Level I | 12 (1.4%) | 17 (2.0%) | 59 (7.0%) | 756 (89.6%) |
| Level II | 4 (0.0%) | 30 (2.1%) | 41 (2.9%) | 1411 (95.0%) |
| Level III | 9 (9.0%) | 7 (7.0%) | 19 (19.0%) | 65 (65.0%) |
| RPC | 8 (17.0%) | 6 (12.8%) | 19 (40.4%) | 14 (29.8%) |
| Abbreviations: RPC, regional perinatal center. |  |  |  |  |

### Objective 3: Characterization of the inter-region maternal transport

**Appendix Table B1** describes the characteristics of inter-region transports. The Macon and Albany DPRs were the most active in inter-region maternal transport. 90% of facilities in Macon and 86% of facilities in Albany transported across DPRs. The Atlanta and Augusta regions were the least active in inter-region transport, with only 50% and 57% of facilities transporting across DPRs, respectively. Overall, 65% of facilities coordinated at least one transport with a facility in another DPR, but inter-region transports account for less than 17% of all transports. 95.1% of inter-region transports occur between facilities located in neighboring regions (**Table 3**).

**Table 3.** Maternal transport by origin and destination DPRs.

|  | *Destination DPR* |  |  |  |  |  |
| --- | --- | --- | --- | --- | --- | --- |
|  | Albany | Atlanta | Augusta | Columbus | Macon | Savannah |
| ***Origin DPR*** |  |  |  |  |  |  |
| Albany | 360 | 2 | 6 | 7 | 11 | 2 |
| Atlanta | 0 | 373 | 6 | 6 | 9 | 1 |
| Augusta | 0 | 5 | 104 | 0 | 0 | 3 |
| Columbus | 41 | 22 | 0 | 258 | 52 | 0 |
| Macon | 185 | 0 | 20 | 26 | 527 | 24 |
| Savannah | 5 | 4 | 20 | 1 | 10 | 667 |

Abbreviations: DPR, designated perinatal region.
a. Grey shading indicates neighboring regions.
b. Number of inter-region transports across neighboring DPRs were 445 out of 468 (95.1%) inter-region transports.

**Table 4** shows factors associated with inter-region transport. Pregnant people were more likely to be transported across DPRs if they had commercial insurance (adjusted OR: 1.418, 95% CI: 1.059-1.900, p = 0.019, ref: Medicaid insured) and if the origin and destination facilities were within the same hospital system (adjusted OR: 1.378, 95% CI: 1.040-1.827, p = 0.026).

**Table 4.**
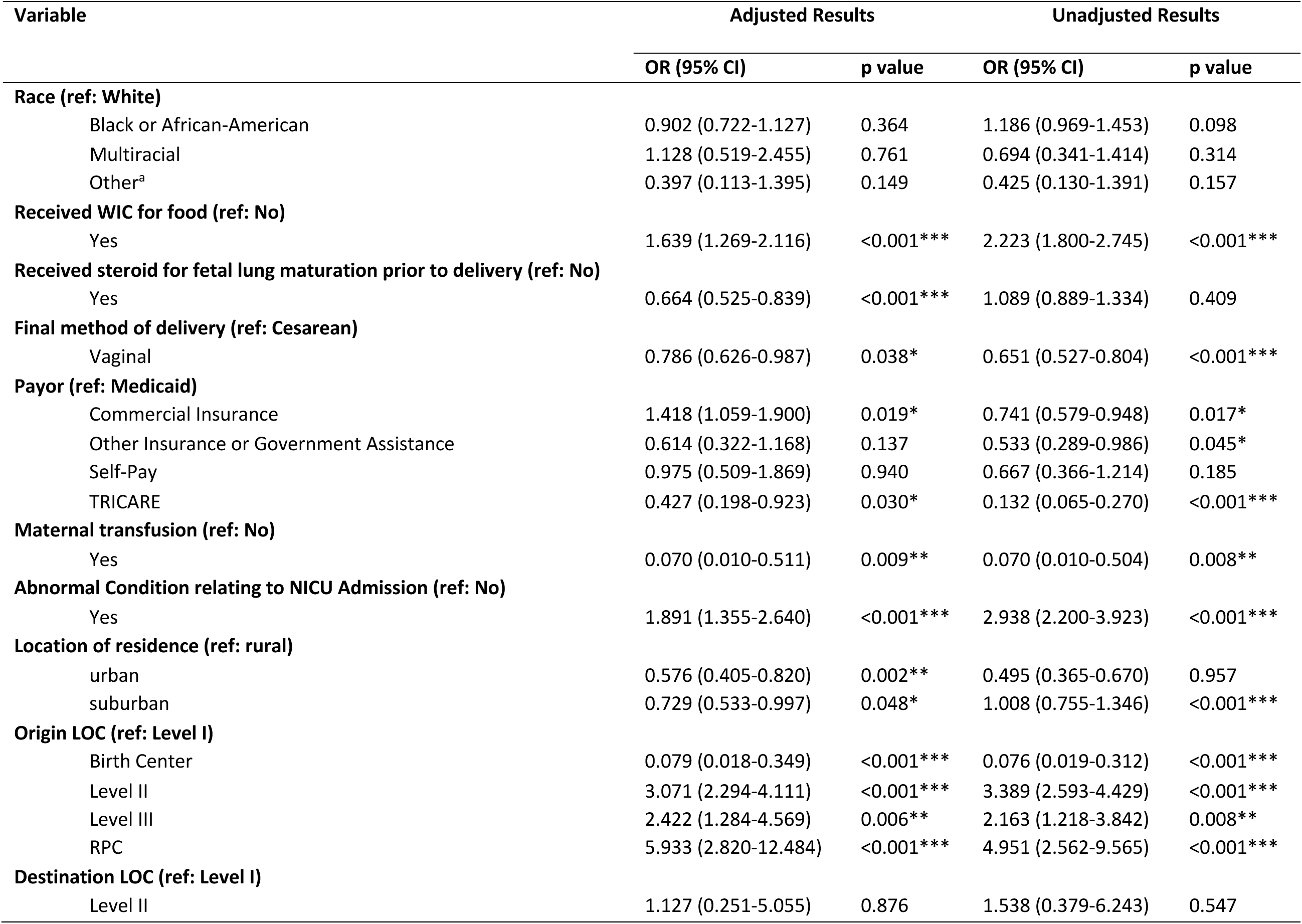
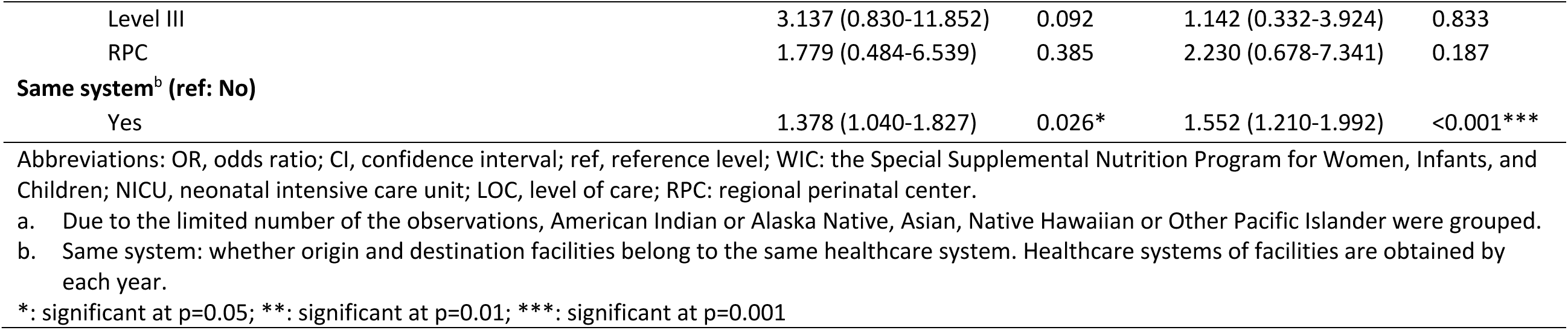
Characteristics associated with inter-region transports.

Subsequent NICU admission (adjusted OR: 1.891, 95% CI: 1.355-2.640, p < 0.001) was also associated with inter-region transports. Conversely, those who received steroid for fetal lung maturation (adjusted OR: 0.664, 95% CI: 0.525-0.839, p < 0.001), had vaginal delivery as a final delivery method (adjusted OR: 0.786, 95% CI: 0.626-0.987, p = 0.038, ref: Cesarean), were on TRICARE (adjusted OR: 0.427, 95% CI: 0.198-0.923, p = 0.030, ref: Medicaid insured), or lived in urban (adjusted OR: 0.576, 95% CI: 0.405-0.820, p = 0.002, ref: rural residence) or suburban areas (adjusted OR: 0.729, 95% CI: 0.533-0.997, p = 0.048, ref: rural residence) were less likely to be transported across DPRs.

### Objective 4: Comparison of observed networks and state guidelines for regionalization

Eight algorithm-detected communities were identified. There were similarities and differences between the state-defined DPRs and algorithm-detected communities (**Table 5**). The Savannah DPR mostly coincided with an algorithm-detected communities where all its facilities were identified in one community with one additional facility in Atlanta DPR (dark green nodes in **Figure 4**). However, the Atlanta DPR consisted of three smaller communities: one consisting of facilities in the northeast of the state (pink), one consisting of a birth center and a Level III facility (gray), and all others in Atlanta DPR (light green). A Chi-square test for independence rejects the null hypothesis that DPRs are not associated with algorithm-detected communities (p < 0.001).

**Figure 4.**
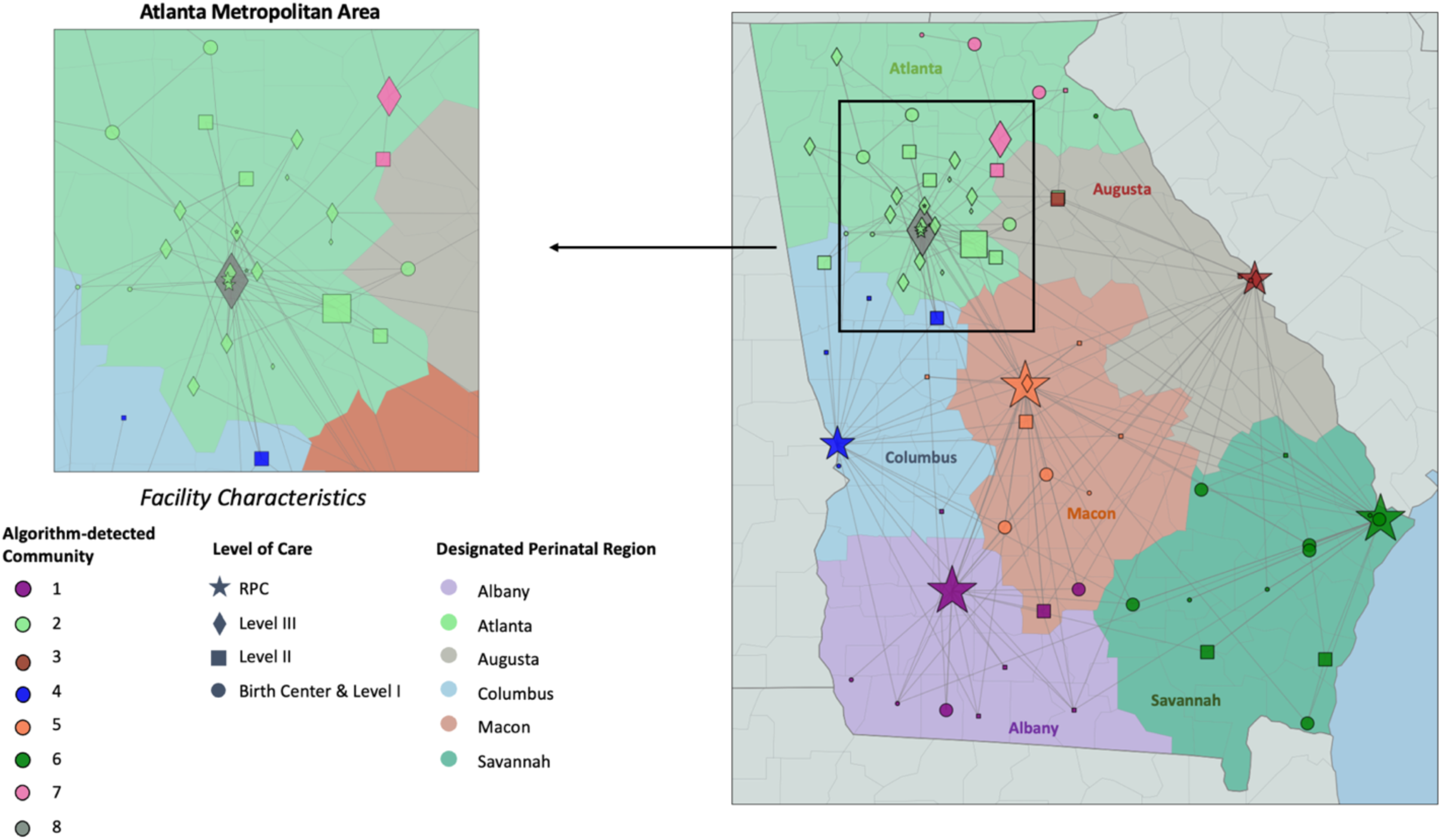
Empirical maternal transport network in Georgia. Shapes of nodes represent levels of care. Colorings of the background regions represent state’s designated regions (DPRs). Colorings of each node represent the algorithm-detected community. Node size is adjusted to be proportional to the incoming transport volume. We omit the arrows indicating the direction of the transport for easier visualization. The Atlanta metropolitan area is zoomed in on the upper left.

**Table 5.** Mismatch between algorithm-detected communities and state-defined DPRs.

| Observed Network | Albany | Atlanta | Augusta | Columbus | Macon | Savannah | Percent Mismatch |
| --- | --- | --- | --- | --- | --- | --- | --- |
| *Algorithm-detected communities* |  |  |  |  |  |  |  |
| **1** | 7 | 0 | 0 | 1 | 2 | 0 | 30.00 |
| **2** | 0 | 25 | 2 | 2 | 0 | 0 | 13.79 |
| **3** | 0 | 0 | 5 | 0 | 0 | 0 | 0 |
| **4** | 0 | 0 | 0 | 5 | 0 | 0 | 0 |
| **5** | 0 | 0 | 0 | 1 | 8 | 0 | 11.11 |
| **6** | 0 | 1 | 0 | 0 | 0 | 13 | 7.14 |
| **7** | 0 | 6 | 0 | 0 | 0 | 0 | 0 |
| **8** | 0 | 2 | 0 | 0 | 0 | 0 | 0 |
| **Percent Mismatch** | 0 | 26.47 | 28.57 | 44.44 | 20 | 0 |  |

a. Chi-square test statistic: 286.34; degree of freedom: 35; $p < 0.001$ ; reject the null hypothesis: there is a significant association between variables, meaning that the state-defined regions are associated with or influenced by the algorithm-detected communities, or vice versa.
b. Dark grey coloring represents the dominated designated perinatal region/detected community for both row and column; light gray coloring represents either the dominated designated perinatal region or detected community.
c. Percent mismatch was calculated by dividing the number of facilities in the non-dominated perinatal region (row)/detected community (column) by the total number of facilities in each detected community (row)/perinatal regions (column).

## Discussion

Our study builds upon prior efforts to characterize perinatal regionalization systems and identify opportunities to re-design these systems to improve maternal outcomes.^11,22^ The most closely related studies to this work are those examining neonatal transport networks in California. ^17,18^ These studies demonstrated that most neonatal transports aligned with the state’s perinatal regions. To our knowledge, our analysis is the first to quantify aspects and dynamics in perinatal regionalization on maternal care using network analysis and is among the few studies that characterize operationalization of maternal transport.^11,13,14,23,24^

There was large county-level variation in maternal transport rates, with multiple counties, particularly in the southeastern Macon DPR and southwestern Albany DPR, exhibiting much higher rates than the state-wide average. We posit that the identification of regions with elevated maternal transport rates could serve as an indicator of a lack of access to risk-appropriate obstetric care because a maternal transport indicates a pregnant person was not immediately able to seek care at a facility that is equipped to handle her pregnancy. This analysis then provides a new view of maternal care access and highlights regions that may be overlooked by other maternal healthcare access metrics. Specifically, two counties with top maternal transport rates (Crisp and Thomas counties) are deemed to have “Full Access” to maternity care according to the March of Dimes despite some pregnant people in these counties living farther than 50 miles from Level III facilities or higher.^25–27^ Thus, the high transport rates in these counties may reveal that current measures over-estimated access to care in Crisp and Thomas counties.

Our results also suggest that transport rates were elevated in suburban and rural counties compared to urban counties. These increased transport rates, especially following the recent closure of rural obstetric units,^28–30^ highlight the need to integrate a rural healthcare perspective into the design of perinatal regionalization guidelines.^24^ Identifying regions with high rates of maternal transport could inform where to best deliver targeted interventions, such as telehealth coverage, collaborative care models, and home visits, to improve access to maternal care.

Further, our results showed significant variation in the characteristics of pregnant people who were involved in a maternal transport. Specifically, pregnant people who were Medicaid- or TRICARE-insured compared to those commercially insured, were more likely to be transported. Previous studies have shown that Medicaid-insured populations make up a disproportionate share of those who live further than 50 miles from Level III or higher care in Georgia.^25^ Conversely, Black pregnant people disproportionately live within 50 miles of Level III or higher care.^25,31^ Thus, geographic distance from higher levels may only be part of the reason that certain populations are transported at higher rates than the general birthing population. The high transport rate among TRICARE-insured pregnant people appears unusual given that TRICARE has largely healthy population and robust coverage.^32^ While the reasons for the high transfer rate among this group are not entirely clear, the elevated transfer rate could be driven by movement between preferred facilities within the TRICARE network rather than medical necessity. Our finding also suggested that complicated deliveries, including preterm birth, multiple birth, and medical conditions of the pregnant person or complications of the infants were associated with maternal transports. These findings are consistent with literature that insurance status and maternal/infant conditions are among the common reasons for maternal transport.^33^ Clinical conditions of pregnant people can further complicate perinatal regionalization by influencing the urgency and complexity of maternal transport. Pregnant people with these conditions may require specialized care that is usually not evenly distributed across facilities, which highlight the need for pre-established yet flexible care coordination systems of maternal transports.

The observed maternal transport network was mostly consistent with Georgia’s designated perinatal regions with a few notable exceptions. First, the community-detection algorithm identified two facilities in the southern part of the Macon DPR which effectively transported as if they were part of the Albany DPR. Second, a group of six facilities in the Northeast part of the Atlanta DPR acted as their own DPR. Furthermore, while inter-region transport was relatively rare (17%), a large proportion of facilities (65%) participated in at least one inter-region transport during the study period. Inter-region transports also disproportionately occurred when the origin and destination facilities were within the same health system. These findings demonstrate that most maternal transports in Georgia occurred within the state’s DPRs, but geographic proximity and transporting to a facility in the same health system tended to trump these designations.

These findings raise questions about whether the design of Georgia’s DPRs should be revisited. Since 2013, the distribution of birthing people in the state of Georgia has changed, and by December 2022, 14 hospitals had closed their obstetric units.^34–36^ Among these was one Level III obstetric facility in Atlanta which ceased operations in November 2022 due to financial challenges and unsustainable operational losses.^37^ Notably, this facility had the highest incoming maternal transport volumes and was a major provider for low-income residents in the city of Atlanta. The closure of such a critical facility further highlights the growing gaps in maternal care across the state.

Redesigning the state’s DPRs to align with the observed empirical networks could formalize relationships that are naturally occurring in practice and optimize care coordination to match current obstetric resources with the current geographic distribution of the birthing population. In Georgia, this could potentially mean the introduction of a new DPR which is consistent with the algorithm-detected community in the Northeast of the Atlanta DPR. If this algorithm-detected community were to be established as a DPR, the Level III facility in Northeast Georgia could serve as an RPC. Moreover, redrawing the Albany DPR to include two other facilities which are 104 miles and 111 miles away from their current RPC in Macon DPR, respectively, would be more consistent with the observed transport patterns and reduce their distances to their newly designated RPC in Albany DPR to 45 miles and 63 miles, respectively.

Geographic distance to care is only one dimension that should be considered when designing perinatal regions. Risk-appropriateness and the patient volume distribution across the system to maintain patient safety and financial stability for facilities may be other important dimensions to consider. The multiple dimensions of a high-performing regionalization system need a more rigorous approach to design perinatal regions based on a set of criteria, such as the process used by the Organ Procurement and Transplantation Network and Trauma Network.^38,39^ Such a process would formalize the criteria that should inform and would prompt changes to the design of perinatal regions as a state’s birthing population distribution shifts and obstetric facilities close.^28–30^ Systems science approaches, such as mathematical optimization and simulation studies, could be used to design regions that would optimally balance these potentially-competing criteria.^40,41^

Transports within the same health system could occur to facilitate care coordination. However, there may be economic reasons for transporting patients within the same system. Reimbursement has been noted as a barrier to regionalization, especially when a maternal transport is involved.^23,42^ As obstetric services are typically billed through global procedure codes at the end of the pregnancy, the origin lower-level facilities might not be reimbursed for any services rendered before the transport, as patients end up delivering at a higher-level facility.^42^ This motivates an examination of outcomes to determine when the potential benefit of care coordination achieved by keeping a patient within the same health system may be outweighed by the increased harm associated with the transport compared to transporting the patient to a closer risk-appropriate facility. If the potential benefits do not outweigh the harms, then a reexamination of reimbursements for transports could be warranted to improve the operationalization of maternal transports. Value-based payment structures may be introduced to incentivize optimal maternal transport.

There are several limitations of our study. First, our study included only deliveries in Georgia, which is a state characterized by a large metropolitan area around the city of Atlanta and extensive rural regions, and thus, our results might not generalize to other states. Our analysis was also limited by the data available in Georgia birth records. We did not have data on out-of-state residents delivering in Georgia or Georgia residents delivering out-of-state. As a result, we were unable to assess care coordination between obstetric facilities in Georgia and obstetric facilities in neighboring states. Additionally, we were not able to characterize risk-appropriateness of the facilities involved in transport due to limited numbers of maternal morbidity indicators available on birth records.

In conclusion, this study is the first to use a network analysis approach to understand the operationalization of maternal transport within an emerging regionalized system of risk-appropriate maternal care. The work found significant variation in the geographic regions and demographic groups that participated in maternal transport. Regions, especially rural counties, with elevated maternal transport rates may lack access to risk-appropriate care. While maternal transports mostly aligned with Georgia’s designated perinatal regions, geography and within health system transport tended to trump these formal designations. Redesigning state’s DPRs to better reflect the observed transport pattern could formalize existing care coordination patterns and match care coordination to current obstetric resources. Future research could explore the economic impacts of maternal transport guidelines, with particular attention to cases involving the transport of patients from Level I facilities to RPCs as opposed to a Level II facility. This analysis could help understand how financial considerations, such as payor policies and reimbursement challenges for back transports, impact these transport decisions.

Additionally, it is beneficial to explore care coordination throughout pregnancy, including how prenatal care and referrals to delivery facilities are coordinated, and their impact on postpartum outcomes. Lastly, linking birth records with administrative data could draw inferences between maternal transport and broader maternal outcomes, and thus understand when maternal transport is warranted.

## Data Availability

The data in this study are not available to be shared by the authors due to our data use agreement.

## Declarations

### Ethics Approval

IRB approval was obtained from the Georgia Institute of Technology (Protocol H23091).

## Funding Information

This study was supported by the Georgia Clinical and Translational Science Alliance and National Center for Advancing Translational Sciences of the National Institutes of Health under Award number UL1TR002378 and the Harold R. and Mary Anne Nash endowment to the Georgia Institute of Technology. The content is solely the responsibility of the authors and does not necessarily represent the official views of the National Institutes of Health.

### Authors’ Contributions

Jingyu Li: conceptualization, data curation, methodology, formal analysis, validation, writing – original draft, visualization.

Stephanie M. Radke: validation, writing – review and editing.

Lauren N. Steimle: data acquisition, conceptualization, methodology, writing – original draft, project administration, funding acquisition, supervision.

## References

1. Hoyert DL. Maternal mortality rates in the United States, 2021. Accessed September 6, 2024. https://stacks.cdc.gov/view/cdc/124678

2. Petersen EE, Davis NL, Goodman D, et al. Racial/Ethnic Disparities in Pregnancy-Related Deaths - United States, 2007-2016. MMWR Morb Mortal Wkly Rep. 2019;68(35):762-765. doi:10.15585/mmwr.mm6835a3

3. CDC. Pregnancy Mortality Surveillance System. Maternal Mortality Prevention. May 20, 2024. Accessed September 5, 2024. https://www.cdc.gov/maternal-mortality/php/pregnancy-mortality-surveillance/index.html

4. Minion SC, Krans EE, Brooks MM, Mendez DD, Haggerty CL. Association of Driving Distance to Maternity Hospitals and Maternal and Perinatal Outcomes. Obstet Gynecol. 2022;140(5):812–819. doi:10.1097/AOG.0000000000004960

5. Kozhimannil KB, Interrante JD, Henning-Smith C, Admon LK. Rural-Urban Differences In Severe Maternal Morbidity And Mortality In The US, 2007-15. Health Aff Proj Hope. 2019;38(12):2077-2085. doi:10.1377/hlthaff.2019.00805

6. Hostetter M, Klein S. Restoring Access to Maternity Care in Rural America | Commonwealth Fund. Accessed July 24, 2024. https://www.commonwealthfund.org/publications/2021/sep/restoring-access-maternity-care-rural-america

7. Ryan GM. Toward improving the outcome of pregnancy Recommendations for the regional development of perinatal health services. Obstet Gynecol. 1975;46(4):375–384.

8. Stark AR, American Academy of Pediatrics Committee on Fetus and Newborn. Levels of neonatal care. Pediatrics. 2004;114(5):1341–1347. doi:10.1542/peds.2004-1697

9. 9. American College of Obstetricians and Gynecologists and Society for Maternal–Fetal Medicine, Menard MK, Kilpatrick S, et al. Levels of maternal care. Am J Obstet Gynecol. 2015;212(3):259-271. doi:10.1016/j.ajog.2014.12.030

10. 10. American Association of Birth Centers;, Association of Women’s Health, Obstetric and Neonatal Nurses;, American College of Obstetricians and Gynecologists, et al. Obstetric Care Consensus #9: Levels of Maternal Care: (Replaces Obstetric Care Consensus Number 2, February 2015). Am J Obstet Gynecol. 2019;221(6):B19-B30. doi:10.1016/j.ajog.2019.05.046

11. DeSisto CL, Oza-Frank R, Goodman D, Conrey E, Shellhaas C. Maternal transport: an opportunity to improve the system of risk-appropriate care. J Perinatol Off J Calif Perinat Assoc. 2021;41(9):2141–2146. doi:10.1038/s41372-021-00935-9

12. Clapp MA, James KE, Kaimal AJ. The effect of hospital acuity on severe maternal morbidity in high-risk patients. Am J Obstet Gynecol. 2018;219(1):111.e1-111.e7. doi:10.1016/j.ajog.2018.04.015

13. Fertaly K, Javorka M, Brown D, Holman C, Nelson M, Glover A. Obstetric transport in rural settings: Referral and transport of pregnant patients in a state without a perinatal regionalized system of care. Health Serv Res. 2024;59(5):e14365. doi:10.1111/1475-6773.14365

14. DeSisto CL, Okoroh EM, Kroelinger CD, Barfield WD. Summary of neonatal and maternal transport and reimbursement policies—a 5-year update. J Perinatol Off J Calif Perinat Assoc. 2022;42(10):1306–1311. doi:10.1038/s41372-022-01389-3

15. DeSisto CL, Kroelinger CD, Levecke M, Akbarali S, Pliska E, Barfield WD. Maternal and neonatal risk-appropriate care: gaps, strategies, and areas for further research. J Perinatol Off J Calif Perinat Assoc. 2023;43(6):817–822. doi:10.1038/s41372-022-01580-6

16. Easter SR, Robinson JN, Menard MK, et al. Potential Effects of Regionalized Maternity Care on U.S. Hospitals. In: Obstetrics and Gynecology. Vol 134. Lippincott Williams and Wilkins; 2019:545-552. doi:10.1097/AOG.0000000000003397

17. Kunz SN, Zupancic JAF, Rigdon J, et al. Network analysis: a novel method for mapping neonatal acute transport patterns in California. J Perinatol Off J Calif Perinat Assoc. 2017;37(6):702–708. doi:10.1038/JP.2017.20

18. Kunz SN, Helkey D, Zitnik M, et al. Quantifying the variation in neonatal transport referral patterns using network analysis. J Perinatol. 2021;41(12):2795–2803. doi:10.1038/s41372-021-01091-w

19. Iwashyna TJ, Christie JD, Kahn JM, Asch DA. Uncharted Paths: Hospital Networks in Critical Care. Chest. 2009;135(3):827–833. doi:10.1378/chest.08-1052

20. Georgia Department of Public Health. Core Requirements and Recommended Guidelines for Designated Regional Perinatal Centers. Georgia Department of Public Health; 2017.

21. Hagberg A, Swart PJ, Schult DA. Exploring Network Structure, Dynamics, and Function Using NetworkX. Los Alamos National Laboratory (LANL), Los Alamos, NM (United States); 2008. Accessed September 7, 2024. https://www.osti.gov/biblio/960616

22. Van Otterloo LR, Connelly CD. Risk-Appropriate Care to Improve Practice and Birth Outcomes. J Obstet Gynecol Neonatal Nurs JOGNN. 2018;47(5):661–672. doi:10.1016/j.jogn.2018.05.004

23. Okoroh E, Kroelinger C, Lasswell S, Goodman D, Williams A, Barfield W. United States and territory policies supporting maternal and neonatal transfer: review of transport and reimbursement. J Perinatol Off J Calif Perinat Assoc. 2016;36(1):30–34. doi:10.1038/jp.2015.109

24. Holman C, Glover A, Fertaly K, Nelson M. Operationalizing risk-appropriate perinatal care in a rural US State: directions for policy and practice. BMC Health Serv Res. 2023;23(1):601. doi:10.1186/s12913-023-09552-y

25. Meredith M, Radke S, Steimle LN. The implications of using maternity care deserts to measure progress in access to obstetric care: a mixed-integer optimization analysis. BMC Health Serv Res. Published online 2024. Accessed July 24, 2024. https://link.springer.com/article/10.1186/s12913-024-11135-4

26. March of Dimes. Nowhere to go: Maternity Care Deserts across the U.S. March of Dimes. 2024. Accessed September 7, 2024. https://www.marchofdimes.org/maternity-care-deserts-report

27. Deloitte United States. March of Dimes Maternity Care Deserts Dashboard. Deloitte United States. Accessed September 7, 2024. https://www2.deloitte.com/us/en/pages/life-sciences-and-health-care/articles/march-of-dimes-maternity-care-deserts-dashboard.html

28. The Chartis Group. Rural America’s OB Deserts Widen in Fallout From Pandemic. The Chartis Group; 2023.

29. Daymude AEC, Daymude JJ, Rochat R. Labor and Delivery Unit Closures in Rural Georgia from 2012 to 2016 and the Impact on Black Women: A Mixed-Methods Investigation. Matern Child Health J. 2022;26(4):796–805. doi:10.1007/s10995-022-03380-y

30. Hung P, Kozhimannil KB, Casey MM, Moscovice IS. Why Are Obstetric Units in Rural Hospitals Closing Their Doors? Health Serv Res. Published online 2016. doi:10.1111/1475-6773.12441

31. Kroelinger CD, Brantley MD, Fuller TR, et al. Geographic access to critical care obstetrics for women of reproductive age by race and ethnicity. Am J Obstet Gynecol. 2021;224(3):304.e1-304.e11. doi:10.1016/j.ajog.2020.08.042

32. Zeiser J. TRICARE is the Health Insurance Employers Need to Know. Accessed September 7, 2024. https://blog.selmanco.com/blog/tricare-is-the-health-insurance-employers-need-to-know-with-tricare-supplement

33. Martin S. Inter-facility maternal transport. 2024. Accessed September 7, 2024. https://www.uptodate.com/contents/inter-facility-maternal-transport

34. Carl Vinson Institute of Government. Births. 2022. Accessed September 8, 2024. https://georgiadata.org/topics/vital-statistics/births

35. Georgia Department of Public Health. OASIS Web Query Tool, Maternal/Child Health. 2024. Accessed September 8, 2024. https://oasis.state.ga.us/

36. Georgia Hospital Association. Georgia Hospital Closure List. 2024. Accessed September 8, 2024. https://www.gha.org/Advocacy

37. Thomas D, Hart A. After a year, questions remain on Atlanta Medical Center’s closure. The Atlanta Journal-Constitution. https://www.ajc.com/news/health-news/after-a-year-questions-remain-on-atlanta-medical-centers-closure/XIHRFL256RFPXCT3ZZAVEUTRFU/. 2023.

38. Organ Procurement and Transplantation Network. Redesign Map of OPTN Regions - OPTN. 2022. Accessed September 7, 2024. https://optn.transplant.hrsa.gov/policies-bylaws/public-comment/redesign-map-of-optn-regions/

39. American College of Surgeons. Regional Trauma Systems: Optimal Elements, Integration, and Assessment. American College of Surgeons; 2008. https://www.facs.org/media/sgue1q5x/regionaltraumasystems.pdf

40. Corley HW, Roberts SD. A Partitioning Problem with Applications in Regional Design. Oper Res. 1972;20(5):1010–1019.

41. Gunantara N. A review of multi-objective optimization: Methods and its applications. Ai Q, ed. Cogent Eng. 2018;5(1):1502242. doi:10.1080/23311916.2018.1502242

42. Safer Birth Initiative. Strategies for Implementation of Regionalized Risk-Appropriate Maternal Care on a National Scale.; 2022. Accessed September 6, 2024. https://saferbirth.org/wp-content/uploads/10-18-2022-FINAL_AIM_HRSA-LoMC-Report.pdf

